# Increased Risk of Autopsy-Proven Pneumonia with Sex, Season and Neurodegenerative Disease

**DOI:** 10.1101/2021.01.07.21249410

**Authors:** Thomas G. Beach, Aryck Russell, Lucia I. Sue, Anthony J. Intorcia, Michael J. Glass, Jessica E. Walker, Richard Arce, Courtney M. Nelson, Tony Hidalgo, Glenn Chiarolanza, Monica Mariner, Alex Scroggins, Joel Pullen, Leslie Souders, Kimberly Sivananthan, Niana Carter, Megan Saxon-LaBelle, Brittany Hoffman, Angelica Garcia, Michael Callan, Brandon E. Fornwalt, Jeremiah Carew, Jessica Filon, Brett Cutler, Jaclyn Papa, Jasmine R. Curry, Javon Oliver, David Shprecher, Alireza Atri, Christine Belden, Holly A. Shill, Erika Driver-Dunckley, Shyamal H. Mehta, Charles H. Adler, Chadwick F. Haarer, Thomas Ruhlen, Maria Torres, Steve Nguyen, Dasan Schmitt, Mary Fietz, Lih-Fen Lue, Douglas G. Walker, Joseph P. Mizgerd, Geidy E. Serrano

## Abstract

There has been a markedly renewed interest in factors associated with pneumonia, a leading cause of death worldwide, due to its frequent concurrence with pandemics of influenza and Covid-19 disease. Reported predisposing factors to both bacterial pneumonia and pandemic viral lower respiratory infections are wintertime occurrence, older age, obesity, pre-existing cardiopulmonary conditions and diabetes. Also implicated are age-related neurodegenerative diseases that cause parkinsonism and dementia. We investigated the prevalence of autopsy-proven pneumonia in the Arizona Study of Aging and Neurodegenerative Disorders (AZSAND), a longitudinal clinicopathological study, between the years 2006 and 2019 and before the beginning of the Covid-19 pandemic. Of 691 subjects dying at advanced ages (mean 83.4), pneumonia was diagnosed postmortem in 343 (49.6%). There were 185 subjects without dementia or parkinsonism while clinicopathological diagnoses for the other subjects included 319 with Alzheimer’s disease dementia, 127 with idiopathic Parkinson’s disease, 72 with dementia with Lewy bodies, 49 with progressive supranuclear palsy and 78 with vascular dementia. Subjects with one or more of these neurodegenerative diseases all had higher pneumonia rates, ranging between 50 and 61%, as compared to those without dementia or parkinsonism (40%). In multivariable logistic regression models, male sex and a non-summer death both had independent contributions (ORs of 1.67 and 1.53) towards the presence of pneumonia at autopsy while the absence of parkinsonism or dementia was a significant negative predictor of pneumonia (OR 0.54). Male sex, dementia and parkinsonism may also be risk factors for Covid-19 pneumonia. The apolipoprotein E4 allele, as well as obesity, chronic obstructive pulmonary disease, diabetes, hypertension, congestive heart failure, cardiomegaly and cigarette smoking history, were not significantly associated with pneumonia, in contradistinction to what has been reported for Covid-19 disease.

## Introduction

There has been a markedly renewed interest in factors associated with pneumonia, a leading cause of death worldwide [1], due to its frequent concurrence with pandemics of influenza [2,3] and Covid-19 disease [4]. In many studies, predisposing factors to both bacterial pneumonia and pandemic viral lower respiratory infections are wintertime occurrence, older age, male sex, obesity, pre-existing cardiopulmonary conditions, and diabetes [5-25]. Also implicated are age-related neurodegenerative diseases that cause parkinsonism and dementia [26-42]. Pneumonia-associated deaths have been estimated to be twice as high in subjects with dementia [43], and the apolipoprotein E4 allele, a genetic risk factor for Alzheimer’s disease (AD), has been reported to be more common in subjects dying with severe Covid-19 disease [44-48]. Pneumonia overall may be much more common as a cause of death in neurodegenerative disease than is apparent from clinically-determined cause of death reporting, as pneumonia is much more likely to be listed as a cause of death when autopsies are done as compared to when autopsies are not done [43,49-52].

We investigated the prevalence of autopsy-proven pneumonia in the Arizona Study of Aging and Neurodegenerative Disorders (AZSAND), a longitudinal, clinicopathological study of normal aging and neurodegenerative diseases [53], between the years 2006 and 2019, before the beginning of the Covid-19 pandemic. Volunteer participants are primarily residents of metropolitan Phoenix, Arizona. The program includes annual clinical assessments of community volunteers by neuropsychologists and subspecialty neurologists, with eventual whole-body autopsies by medically-certified anatomical pathologists and neuropathologist. The current study was planned to determine the relative frequency of pneumonia in autopsied participants with neuropathologically classified forms of dementia and parkinsonism relative to those who were free of these during life, and to test other possible associations including with sex, age, chronic medical comorbidities and seasonality.

## Methods

### Study subjects and clinical assessments

The study population draws primarily on the retirement communities of greater Phoenix, Arizona. The residents are predominantly elderly, well-educated, Caucasian (greater than 90%), middle- and upper-income individuals. Recruitment is directed at subjects with a clinical diagnosis of Alzheimer’s disease (AD) or Parkinson’s disease (PD) or who are neurologically normal. Most of the clinically-followed subjects are living independently at the time of enrollment, without cognitive impairment or parkinsonism. Subjects with hazardous infectious diseases including HIV, hepatitis B or C, Creutzfeldt-Jakob disease and other infectious encephalopathies are excluded from enrollment.

The AZSAND, through its Brain and Body Donation Program, performs constitutively-rapid autopsies (median postmortem interval 3.0 hours for all 2,000 + autopsies) on Program participants [53]. The Program is approved by an Institutional Review Board and written informed consent is obtained from all subjects or their legal representatives. Tissue and other biospecimens are available to researchers worldwide through our website (https://www.brainandbodydonationregistration.org/).

Standardized general medical, subspecialty neurological (behavioral/cognitive and movement disorders), and neuropsychological assessments are administered to most subjects annually. Clinical diagnostic classification is performed annually and after death, the latter while blinded to autopsy findings, at a consensus conference attended by neurologists and neuropsychologists.

Additionally, private medical records are obtained for all subjects and reviewed in a standardized manner. Records spanning the most recent two years are obtained at the time of enrollment and again at the time of death. The records receive multiple reviews by trained clinical research, medical and nursing staff, including a neuropathologist. Subjects also fill out medical history questionnaires at their annual clinical assessment visits.

### Autopsy examinations

Medically-certified pathologists performed all examinations. Lung sampling was directed at both grossly-normal and grossly-pathological regions. The database postmortem diagnoses were queried for “pneumonia”. Acute pneumonia was defined based on microscopic findings, in hematoxylin and eosin-stained slides, of contiguous intra-alveolar accumulations of neutrophil polymorphonuclear leukocytes, with or without a typical gross pathological appearance or microscopic evidence of bacterial colonies. The neuropathological diagnostic approach has been previously described [53]. Published clinicopathological consensus criteria [54-61] were used when applicable, incorporating clinical assessment results as well as pertinent private medical history. Parkinson’s disease (PD) was diagnosed if the subject had two of three cardinal clinical signs in conjunction with Lewy bodies and pigmented neuron loss in the substantia nigra (57). Alzheimer’s disease (AD) was diagnosed when dementia was present during life and there were intermediate or high AD neuropathological changes at autopsy [59].

### Associations tested and statistical methods

Fisher’s Exact Tests were used to test for univariable associations of clinically and autopsy-derived subject characteristics with pneumonia diagnosed at autopsy. Tested characteristics included older age (greater than the median age of 84), sex, season at time of death (summer months of June, July or August versus non-summer months), obesity (defined as body mass index of 30 or greater), chronic obstructive pulmonary disease (COPD), diabetes, hypertension, congestive heart failure, cigarette smoking history, cardiomegaly (autopsied heart weight of > 360 g for males and > 280 g for females) and presence or absence of neuropathologically diagnosed cerebrovascular disease (vascular dementia; VaD) or neurodegenerative disorders including AD, PD, dementia with Lewy bodies (DLB) and progressive supranuclear palsy (PSP). Additionally, hetero- and homozygosity for the ε4 allele was tested for association. Variables with significant (p < 0.05) associations with autopsy-diagnosed pneumonia were further tested for their independent contributions using multivariable logistic regression.

## Results

Subjects all died between the years 2006 and 2019 and before the beginning of the Covid-19 pandemic. There were 691 subjects with postmortem lung examinations (Table 1). Of these, 405 were males (59%) and 286 were females (41%) with mean ages of 82.1 years for males and 85.2 for females. Pneumonia was diagnosed in 343 subjects (49.6%; 225 males and 118 females). This was almost entirely acute bronchopneumonia with only one acute lobar pneumonia, one with “chronic pneumonia”, and one with “non-specific chronic pneumonitis”. Three cases had combined acute and chronic pneumonia or pneumonitis. Clinical and clinicopathological brain diagnoses included 185 without dementia or parkinsonism, 319 with AD, 127 with idiopathic PD, 72 subjects with DLB, 49 with PSP and 78 with VaD. These diagnoses are not mutually exclusive as many of the subjects had more than one neuropathological diagnosis [60]. Of the 319 AD cases, 34 also had clinicopathologically-diagnosed PD, 62 had DLB, 60 had VaD and 16 had PSP. Other neurodegenerative conditions diagnosed but not statistically tested for associations with pneumonia due to low subject numbers included frontotemporal lobar degeneration with TDP-43 proteinopathy (n = 14), corticobasal degeneration (n = 7) and Pick’s disease (n = 4).

**Table 1.** Neuropathological and clinical classification of study subjects. Norm = non-demented without parkinsonism; AD = Alzheimer’s disease; PD = Parkinson’s disease; DLB = dementia with Lewy bodies; VaD = vascular dementia; PSP = progressive supranuclear palsy.

|  | ALL | Norm | AD | PD | DLB | VaD | PSP |
| --- | --- | --- | --- | --- | --- | --- | --- |
| **Number** | 691 | 185 | 319 | 127 | 72 | 78 | 49 |
| **Age (SD)** | 83.4 (9.6) | 85 (3.5) | 84.3 (2.8) | 80 (2.1) | 83.6 (9.2) | 89.3 (10.6) | 86.4 (6.8) |
| **M/F** | 405/286 | 112/73 | 182/137 | 89/38 | 51/21 | 39/39 | 30/19 |

Univariable comparison of subject characteristics in those with and without pneumonia identified male sex, non-summer death and AD and DLB as all being significantly more likely to have had an autopsy diagnosis of pneumonia (Table 2). Female subjects and those without dementia or parkinsonism were significantly less likely to have had an autopsy pneumonia diagnosis. Hetero- or homozygosity for the apolipoprotein E-ε4 allele was not associated with pneumonia, nor were older age, VaD, obesity, diabetes, hypertension, congestive heart failure, cardiomegaly or cigarette smoking history.

**Table 2.** Comparison of subject characteristics of those with and without pneumonia. Fisher Exact tests were used to compare proportions. Obesity is defined as BMI 30 or greater. Cardiomegaly is defined as heart weight > 360 g for males and > 280 g for females. Older age is defined as greater than the median age of 84. Any ApoE4 = apolipoprotein genotype heterozygous or homozygous for the ε4 allele; ApoE 4/4 = homozygous for ε4 allele.

| Variable | Pneumonia | No Pneumonia | Statistic |
| --- | --- | --- | --- |
| Male | 225/343 (65%) | 181/348 (52%) | p = 0.0004 |
| Female | 118/343 (34%) | 168/348 (48%) | P = 0.0006 |
| Older Age | 158/343 (46%) | 178/348 (51%) | ns |
| No Dementia or Parkinsonism | 70/343 (20%) | 115/348 (33%) | p = 0.0002 |
| AD | 172/343 (50%) | 147/348 (42%) | p = 0.04 |
| PD | 72/343 (21%) | 55/348 (16%) | P = 0.09 |
| DLB | 44/343 (13%) | 28/348 (8%) | p = 0.046 |
| VaD | 39/343 (11%) | 39/348 (11%) | ns |
| PSP | 30/343 (9%) | 19/348 (5%) | P = 0.10 |
| Any ApoE4<sup>1</sup> | 113/336 (34%) | 101/345 (29%) | ns |
| ApoE 4/4 | 16/336 (4.8%) | 14/345 (4.6%) | ns |
| Non-Summer Death | 276/343 (80%) | 256/348 (73%) | 0.037 |
| COPD | 78/343 (23%) | 107/348 (31%) | 0.020 |
| Obesity (BMI 30 or greater)<sup>2</sup> | 87/342 (25%) | 92/334 (26%) | ns |
| Cardiomegaly<sup>3</sup> | 252/342 (74%) | 263/343 (77%) | ns |
| Congestive Heart Failure | 74/343 (22%) | 94/348 (27%) | ns |
| Hypertension | 232/343 (68%) | 262/348 (85%) | 0.10 |
| Diabetes | 80/343 (23%) | 84/348 (24%) | ns |
| Cigarette Smoker | 153/343 (45%) | 161/348 (46%) | ns |

1. Ten subjects had no apolipoprotein E genotyping available. 2. Fourteen subjects had no BMI values available. 3. Six subjects had no heart weight available.

With a targeted comparison of pneumonia rates in neurodegenerative and cerebrovascular disease versus subjects without clinically diagnosed dementia or parkinsonism (Table 3), AD, PD, DLB and PSP all had significantly higher rates. The prevalence of pneumonia in subjects without dementia or parkinsonism was 40% and ranged between 50% and 61% in the tested neuropathological disease categories.

**Table 3.** Comparison of prevalences of pneumonia at autopsy in neurodegenerative conditions and vascular dementia versus subjects without clinically-diagnosed dementia or parkinsonism.

| **Diagnosis** | **Pneumonia** | **Statistic** |
| --- | --- | --- |
| No Dementia or Parkinsonism | 70/185 (40%) | n/a |
| Alzheimer's Disease | 172/319 (54%) | 0.0006 |
| Parkinson's Disease | 72/127 (57%) | 0.0012 |
| Dementia with Lewy Bodies | 44/72 (61%) | 0.0012 |
| Vascular Dementia | 39/78 (50%) | 0.08 |
| Progressive Supranuclear Palsy | 30/49 (61%) | 0.0054 |

Subject characteristics with a significant pneumonia association on univariable analysis (Table 2) were entered into multivariable logistic regression models. Only sex, absence of parkinsonism or dementia and season of death retained significance in the final model (Table 4). Male sex and a non-summer death both had significant independent contributions (ORs of 1.67 and 1.53) towards the presence of acute pneumonia at autopsy while the absence of parkinsonism or dementia was a significant contributor towards the absence of pneumonia (OR 0.54 for pneumonia).

**Table 4.** Final logistic regression model showing subject characteristics retaining independent significance as predictors of the presence of acute pneumonia at autopsy.

| **Predictor** | **Odds Ratio** | **95% Conf. Interval** | **Significance** |
| --- | --- | --- | --- |
| Male Sex | 1.67 | 1.22 – 2.28 | 0.0013 |
| No Dementia Or Parkinsonism | 0.54 | 0.38 – 0.77 | 0.00061 |
| Non-Summer Death | 1.53 | 1.07 – 2.20 | 0.021 |

## Discussion

Although pneumonia is among the leading causes of morbidity and mortality worldwide, we still do not have a comprehensive understanding of host susceptibility factors [62]. Accentuating the urgency for more knowledge are the recent pandemics caused by H1N1 influenza and SARS-CoV-2, which frequently result in concurrent viral and bacterial pneumonia [2-4]. There is evidence that viral and bacterial pneumonia share host vulnerabilities including winter season, older age, male sex, obesity, pre-existing cardiopulmonary conditions, and diabetes [6-25]. Older people with neurodegenerative diseases are also more at risk [26-43], and the apolipoprotein E4 allele, a genetic risk factor for Alzheimer’s disease (AD), has been reported to be more common in subjects dying with severe Covid-19 disease [44-48].

Autopsies provide more accurate diagnoses for both pneumonia and neurodegenerative diseases [43,49-52,63-65]. Nevertheless, even in hospitalized or autopsied patients, a positive identification of responsible pneumonia pathogens may be achieved only in a minority of cases [66]. When pathogens are identified, they are often both viral and bacterial, with many mixed viral and bacterial attributions. The top three pathogens in US hospitalized patients have been reported to be human rhinovirus, influenza A and B, and Streptococcus pneumoniae [67].

Although we did not attempt to determine, from clinical records or postmortem methods, the presence of specific associated pathogens in our study subjects, the contiguous intra-alveolar accumulation of neutrophils is generally regarded as indicative of a bacterial cause [68] and therefore this study is likely to have been biased towards the inclusion of bacterial pneumonia and against the inclusion of purely viral pneumonia, although mixed bacterial and viral causes were very likely to have been common [66-68].

In this study of a suburban-dwelling, elderly and predominantly Caucasian middle-class population, male sex and non-summer deaths were roughly equivalent as predictors of autopsy-proven acute pneumonia, with ORs of 1.67 and 1.53, respectively. The absence of clinically-documented parkinsonism or dementia was associated with significantly less likelihood of pneumonia at death, with an OR of 0.54 for the presence of pneumonia.

The strong association with male sex has been previously reported, not only for pneumonia [16,69,70], but also for sepsis [71], but not without opposing evidence [72,73]. Covid-19-associated pneumonia has a strong proclivity for males [11-13,15,18,23]. Suggested reasons for increased pneumonia in males have included a disparity in the immunosuppressive vs immunostimulatory effects of male versus female sex steroids or in the balance of circulatory pro-inflammatory versus anti-inflammatory mediators [71], as well as relatively less effective humoral and cellular immunity responses in men [23]. We considered whether the male predominance of PD and PSP may have influenced the overall male pneumonia association and therefore we repeated the final logistic regression model after excluding all PD and PSP cases; this did not appreciably change the ORs or their significances (results not shown).

The wintertime association with pneumonia has been repeatedly documented and is likely due to the predominant winter occurrence of influenza [74,75].

Multiple studies have reported that pneumonia is more likely to occur together with dementia and other neurodegenerative diseases [26-42]. Attems et al [26], in an autopsy study of 308 hospitalized patients, found bronchopneumonia to be the cause of death in 45% and 28% of those with and without dementia, a significant difference, but neuropathologically-defined dementia subsets, including AD, VaD, DLB and PSP, were not statistically different in their pneumonia rates. Brunstrom and Englund [27] studied 524 hospitalized and autopsied patients with dementia, finding bronchopneumonia to be the cause of death in 38% versus only 3% in the general (non-autopsied) population and significantly more common in AD vs VaD (47% vs 27%). Magaki et al [38] found pneumonia as the cause of death in 66% of 86 autopsied dementia cases, as compared with 21% in 124 autopsies of non-demented subjects. Subtypes of dementia were not sufficient for statistical analysis but showed a trend for much higher pneumonia rates in AD as compared to VaD, DLB, PD and PSP. A death certificate study by Hobson and Meara [31] found pneumonia as a significantly greater cause of death in PD vs non-PD (53% vs 21%; OR = 2.0) while Fall et al [37] had findings in the same direction (24% vs 8%; hazard ratio = 7.5). Litvan et al [29] found pneumonia to be the cause of death in 13/20 (65%) of patients with PSP while for Nath et al [42] this was 22/49 (45%). Manabe et al [40] reported, for subjects with neuropathologically confirmed DLB, pneumonia as the clinically identified cause of death in 39/42 (93%), and a meta-analysis [43] of clinical and autopsy studies estimated the OR for a pneumonia death as 2.0 for all-cause dementia vs no dementia. A meta-analysis of 13 mixed clinical and autopsy studies by Foley et al [39] found pneumonia death ORs of 2.2 for any dementia vs 1.7 for AD specifically.

The results of the current study advance this field by providing superior pneumonia and brain pathology diagnoses, as compared to clinical or death certificate attribution alone, by the performance of whole-body autopsies with relatively large subject numbers and by simultaneously considering the relative contributions of other possible factors including sex, age, season of death, neuropathologically-defined subtypes for both dementia and parkinsonism, cardiopulmonary conditions and possession of the apoE4 allele. This is the first study that we know of that has found male sex to be an independent pneumonia risk factor when considered together with neurodegenerative disease. Subjects without dementia or parkinsonism were only half as likely to die with pneumonia, confirming prior studies. In our study, subtypes of parkinsonism and dementia did not retain significance on multivariable logistic regression but collectively are implicated by the strong negative association of pneumonia with the absence of these. All tested neurological conditions except VaD were significant on univariable comparisons with non-demented, non-parkinsonian controls. The apoE4 allele was not more common, in the current study, in subjects dying with pneumonia, which may point to an important difference with Covid-19 disease, where the apoE4 allele has been reported as a significant association [44-48].

Surprisingly, older age and all of the cardiopulmonary diseases and risk factors analyzed were not associated with increased rates of pneumonia. A large study, across all ages, of community-acquired pneumonia requiring hospitalization, found a markedly increased incidence among those aged 80 or over [67]. It is likely that the generally advanced ages of our entire study group did not allow for a distinction based on relative age. We considered the possibility that the high prevalence of neurodegenerative disease in our study population may have resulted in a statistically dominating influence on the presence or absence of pneumonia, thereby obscuring associations with the cardiopulmonary conditions that other studies have found to be pneumonia risk factors. To explore this possibility, we repeated the univariable analyses in the subset of subjects without a diagnosis of parkinsonism or dementia (n = 185).

None of the comparisons (results not shown) were significant but again there was a trend for a greater likelihood of pneumonia in males (p = 0.17) and pneumonia was again more common with non-summer deaths (p = 0.10). Obesity, COPD, cardiomegaly, congestive heart failure, hypertension, diabetes and cigarette smoking were not statistically associated with pneumonia. Standardized clinical criteria were not used to clinically define these comorbid conditions and thus there may have been misdiagnoses. Obesity was defined by body mass index and cardiomegaly by weight at autopsy, giving more confidence for these. There is a suggestion, in fact, of a paradoxically decreased prevalence of typical cardiovascular risk factors and disease in our pneumonia subjects, with rates of several investigated factors being lower than in those without pneumonia (see Table 2). This may be due to the lower prevalences of cardiovascular diseases and associated risk factors in AD and PD [76,77] and perhaps other neurodegenerative disorders, with a resultant tendency to segregate in those without these disorders.

We conjecture, as have others, that increased rates of pneumonia in subjects with an advanced neurodegenerative disease may be due in large part to neurological impairment of swallowing with resultant greater risk of aspiration [39, 78] but there are likely also contributory social or cognitive and behavioral factors. These include lack of awareness, attention and executive control, and impulsivity [78]. Individuals with AD and other neurodegenerative disorders are more likely to reside in communal, assisted living facilities, with associated increased risk of infections, influenza and pneumonia, and a tendency to become bedridden in their last months of life. They may be less able to communicate their symptoms to caregivers and less likely to receive effective treatment. As many of our subjects were on hospice care, they were more likely to be receiving medications such as anti-psychotics, narcotics and benzodiazepines that may increase aspiration risk, and they were more likely to have had advance directives including “do not intubate”, or which precluded the use of antibiotics.

We did not investigate whether other acute or chronic conditions, such as metastatic cancer, acute myocardial infarction or stroke, might have influenced the occurrence of acute pneumonia, and it is possible that any of these may have been additional significant factors.

We did not conduct detailed morphological or immunophenotyping analyses of lungs with and without pneumonia. This will be an important future objective.

Non-Covid-19 pneumonia, like Covid-19 pneumonia, is significantly more likely to occur in males. Published data so far also indicates that people with dementia or parkinsonism are also more susceptible to severe Covid-19 pneumonia [79-86]. The apolipoprotein E4 allele, as well as obesity, chronic obstructive pulmonary disease, diabetes, hypertension, congestive heart failure, cardiomegaly and cigarette smoking history, were not significantly associated with non-Covid-19 pneumonia, in contradistinction to what has been reported for Covid-19 disease.

## Data Availability

All data is available on request to

## Acknowledgements

This project was supported by a Supplement to a National Institute on Aging grant (3P30AG019610-20S1), submitted in response to a Notice of Special Interest (NOSI) issued by the National Institute on Aging (NOT-AG-20-022), “to highlight the urgent need for research on Coronavirus Disease 2019…”. The Brain and Body Donation Program has been supported by the National Institute of Neurological Disorders and Stroke (U24 NS072026 National Brain and Tissue Resource for Parkinson’s Disease and Related Disorders), the National Institute on Aging (P30 AG19610 Arizona Alzheimer’s Disease Core Center), the Arizona Department of Health Services (contract 211002, Arizona Alzheimer’s Research Center), the Arizona Biomedical Research Commission (contracts 4001, 0011, 05-901 and 1001 to the Arizona Parkinson’s Disease Consortium) and the Michael J. Fox Foundation for Parkinson’s Research. JPM is supported by NIH grants R35 HL135756 and R33 HL137081. We acknowledge the contributing technical expertise of Jonette Henry-Watson.

## Notes

### Competing Interest Statement

The authors have declared no competing interest.

### Funding Statement

This project was supported by a Covid-19 Supplement to a National Institute on Aging grant (3P30AG019610-20S1). The Brain and Body Donation Program has been supported by the National Institute of Neurological Disorders and Stroke (U24 NS072026 National Brain and Tissue Resource for Parkinson Disease and Related Disorders), the National Institute on Aging (P30 AG19610 Arizona Alzheimer Disease Core Center), the Arizona Department of Health Services (contract 211002, Arizona Alzheimer Research Center), the Arizona Biomedical Research Commission (contracts 4001, 0011, 05-901 and 1001 to the Arizona Parkinson Disease Consortium) and the Michael J. Fox Foundation for Parkinson Research. JPM is supported by NIH grants R35 HL135756 and R33 HL137081. Otherwise, no authors received payment or services from a third party for any aspect of the submitted work.

### Author Declarations

Western Institutional Review Board, Puyallup, WA, USA

